# Total and Stroke Related Imaging Utilization Patterns During the COVID-19 Pandemic

**DOI:** 10.1101/2020.05.20.20078915

**Authors:** Long H. Tu, Richa Sharma, Ajay Malhotra, Joseph L. Schindler, Howard P. Forman

**Affiliations:** Yale Department of Radiology and Biomedical imaging, Yale University School of Medicine, 330 Cedar St, TE 2-214, New Haven, CT 06520; Yale Department of Neurology, Yale University School of Medicine, 15 York St S, New Haven, CT 06510

## Abstract

During the COVID-19 pandemic, radiology practices are reporting a decrease in imaging volumes. We review total imaging volume, CTA head and neck volume, critical results rate, and stroke intervention rates before and during the COVID-19 pandemic. Total imaging volume as well as CTA head and neck imaging fell approximately 60% since the beginning of the pandemic. Critical results fell 60-70% for total imaging as well as for CTA head and neck. Compared to the same time frame a year prior, the number of stroke codes at the early impact of the pandemic had decreased approximately 50%. Proportional reductions in total imaging volume, stroke-related imaging, and associated critical result reports during the COVID-19 pandemic raise concern for missed stroke diagnoses in our population.

## Introduction

In the midst of the COVID-19 pandemic, medical providers are seeing a decrease in patients seeking care for emergency conditions such as heart attacks and strokes. This was first reported on social and news media [1, 2]. Early hospital-level evidence also supports this observation [3, 4].

Radiology practices are reporting a concurrent decrease in imaging volumes of 50-70% [5, 6]. It is unclear to what extent factors such as patient self-selection, avoidance of necessary care, or changes in utilization contribute to this decrease. The incidence of some emergent medical conditions, such as stroke, is not expected to be directly impacted by infection prevention measures.

We obtained clinical and imaging data at a large, tertiary academic medical center during the COVID-19 pandemic and compared it with stroke data from the corresponding time period from 2019 to gain insight into the contributions of these various factors to imaging volume and critical result rate.

## Materials and Methods

This study was approved for retrospective review by the institutional IRB and patient consent was waived. Review of imaging was performed from 1/1/2020 – 4/13/2020, spanning before and after the impact of COVID-19. Total imaging volume per day during this period was extracted and queried for critical results using key terms in institutional policy. Specific query for computed tomographic angiography (CTA) imaging of the head and head and related critical results was also obtained during the same time period. Chart review of patients with critical results on CTA head and neck was performed to ascertain the detected acute pathology. For each of the above, 7-day rolling averages were calculated.

## Results

Total imaging volume decreased approximately 60% in the study period time frame. At the end of the study period, the 7-day average of studies per day was 916 (on 4/13/2020), compared to 2462 (on 1/13/2020). A sharp change was seen concurrent with response measures for COVID-19 starting in mid-March, beginning on 3/16/2020. Total critical result rates also decreased to an approximately proportionally degree, 60-70% (Figure 1). At the end of the study period, the 7-day average of critical results per day was 70 (on 4/13/2020), compared to 209 (on 1/13/2020). CTAs performed also declined approximately 60% in volume during this time frame. At the end of the study period, the 7-day average was 3.3 (on 4/13/2020), compared to 10.7 (on 1/13/2020). The exact magnitude of decrease is difficult to describe given variability in CTAs performed on a day-to-day and week-to-week basis. The rate of critical results and ischemic pathologies found on imaging is below usual trends, with similar decreases in magnitude (Figure 2). The exact decrease in these figures is similarly difficult to quantify. The imaging volumes have remained at the lower numbers since the mid-March decrease. The number of stroke codes showed a roughly 50% decrease in the initial week of the pandemic compared to the corresponding period in 2019, with even greater decrease in number of patients getting intravenous thrombolysis (IVT) and endovascular thrombectomy (EVT). However, the number of stroke codes and IVT administrations in the subsequent three weeks have increased but remain low compared to the 2019 numbers. The number of EVTs has begun to increase from this low point.

**Figure 1:**
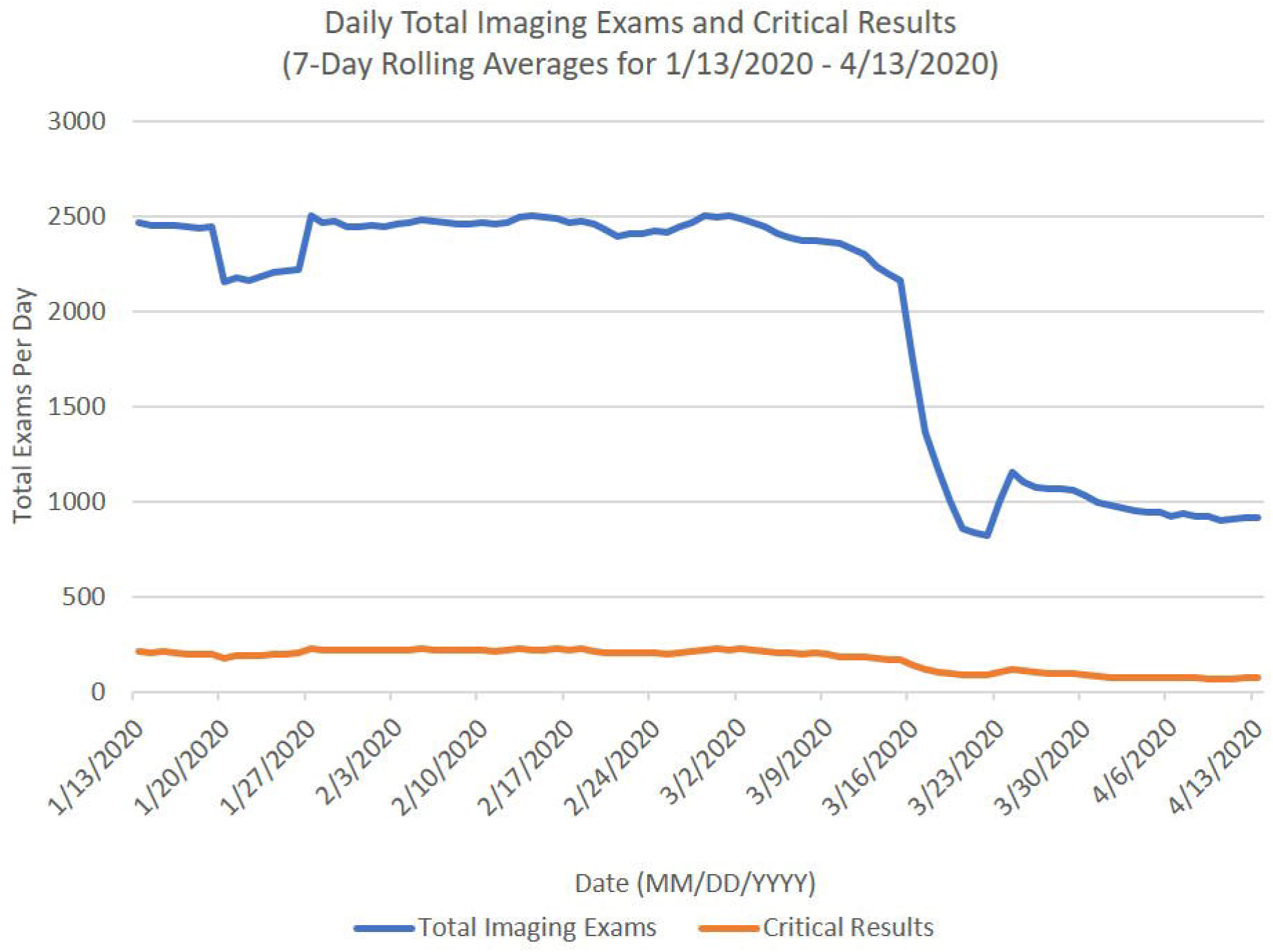
Daily total imaging volume at our institution decreased approximately 60%, from a range of 2400-2500 studies per day to 900-100 studies per day after the impact of COVID-19. Critical results in imaging reports decreased 60-70%, from a range of 200-220 per day to 70-75 per day. The sharp decrease between 3/16/2020 and 3/23/2020 coincides with major institutional response measures and implementation of social distancing guidelines by the CDC.

**Figure 2:**
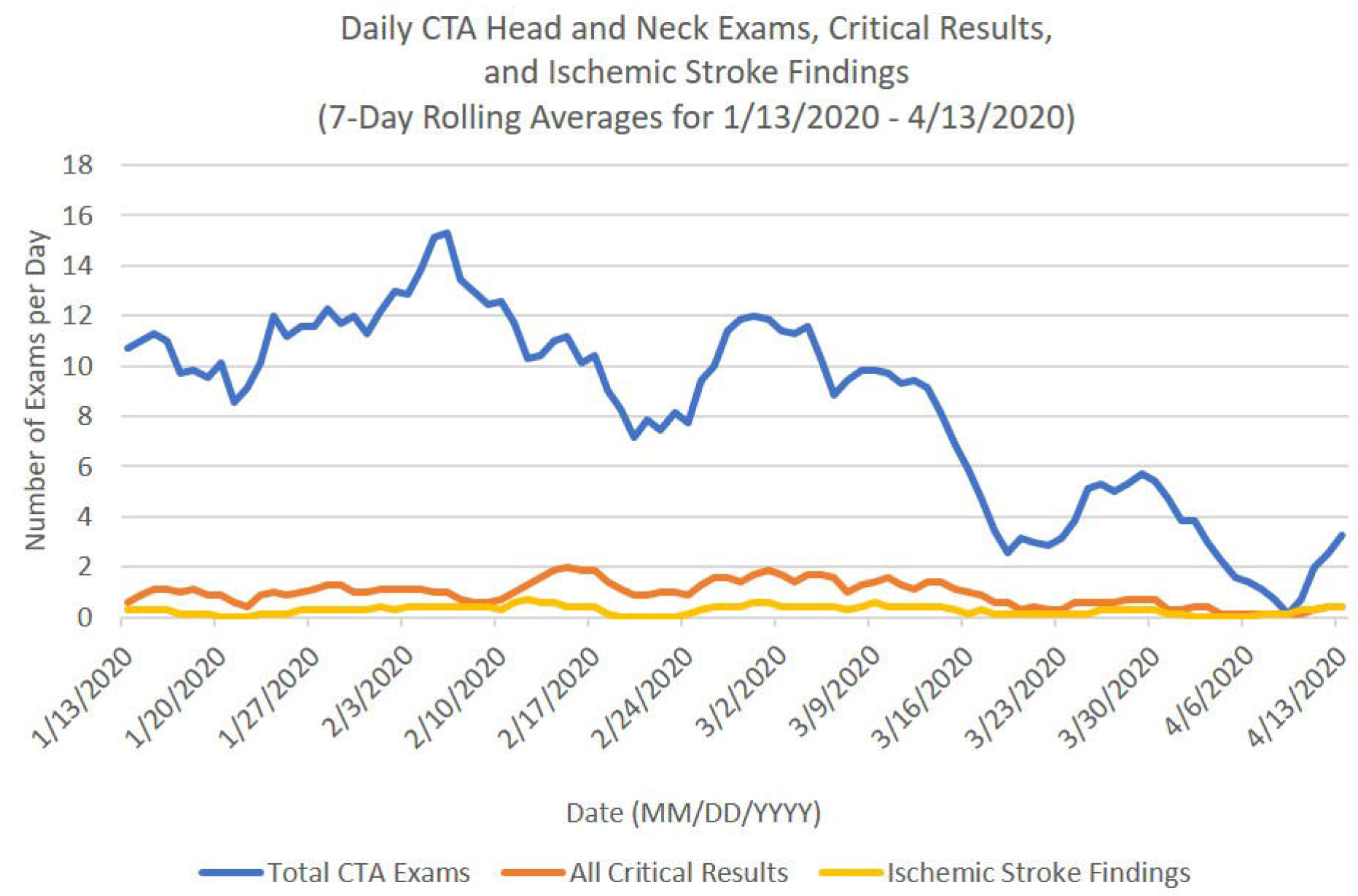
Total CTA head and neck imaging at our institution decreased from a range of 8-15 studies per day to 06 studies per day. Critical results and ischemic pathologies are seen at a decreased rate compared to before the week of 3/16/2020 - 3/23/2020. Critical results in late March and early April range from 0-0.5 per day for 7-day rolling averages. Pre-COVID, this was 1-2 critical results per day. Comparison of ischemic pathologies before and during COVID-19 is difficult due to the low numbers, though follows a similar trend as overall critical results, reaching minimums not otherwise seen in 2020.

## Discussion

Our data demonstrate an approximate decrease of 60% total imaging volume during the COVID-19 pandemic. Total critical result numbers have also decreased by 60-70%. A large proportion of this change may reflect postponement of outpatient imaging studies and exams of ultimately non-emergent pathologies. The proportional decrease in critical results however suggests that acute pathologies in our population may not be presenting to medical attention. There is a similar trend when looking at only CTA head and neck exams with regard to total volume, critical results, and ischemic pathologies. There was an initial dramatic decrease in the stroke codes in the first week of the crisis, with increased numbers since, although still lower compared to 2019 volumes.

Reductions in imaging volume, stroke related findings, and stroke codes during the COVID-19 pandemic raise concern for missed stroke diagnoses, a worrisome possibility. Further work is necessary to assess the long-term impact of this trend.

## Data Availability

Data are available upon reasonable request.

